# Age and sex differences in the modification of the leg lean mass–bone mineral density association by sedentary time in U.S. adults

**DOI:** 10.64898/2026.09.13.26362934

**Authors:** Yasushi Kishimoto

## Abstract

**Background:** Lean mass is positively associated with bone mineral density (BMD), and this relationship may vary according to age and sex. Sedentary behavior has also been associated with BMD, although findings differ across populations and skeletal sites. However, it remains unclear whether sedentary time modifies the relationship between regional lean mass and BMD and whether such modification varies across sex and age. These relationships were explored using nationally representative U.S. data.

**Methods:** Adults aged 18-59 years from the 2017-2018 National Health and Nutrition Examination Survey (NHANES) were analyzed. Mean bilateral leg lean mass and mean bilateral leg BMD were derived from dual-energy X-ray absorptiometry measurements, and sedentary time was obtained from self-report. Survey-weighted regression models incorporated NHANES examination weights, strata, and primary sampling units. The primary model examined the four-way interaction among leg lean mass, sedentary time, sex, and continuous age, with adjustment for race/ethnicity and height. Quadratic lean-mass terms were examined in sensitivity analyses.

**Results:** The primary complete-case analysis included 2,543 adults (1,191 men and 1,352 women). Women showed a more apparent decline in both leg lean mass and leg BMD at ages 50-59 years, whereas sedentary time did not increase in parallel with these age-related changes. Leg lean mass was positively associated with leg BMD in both sexes, with modest nonlinearity, particularly among women. In the primary survey-weighted model, the four-way interaction among leg lean mass, sedentary time, sex, and age was positive (beta = 0.000111 per year, 95% CI 0.000039-0.000183; P = 0.0051), indicating increasing sex divergence in sedentary-time modification of the lean mass-BMD association with age. At younger ages, conditional interaction estimates were similar between sexes; at older ages, estimates became increasingly positive in women while remaining negative but imprecise in men. Quadratic sensitivity analyses attenuated the four-way interaction estimate by approximately 9-12% and reduced its statistical precision, while broadly preserving its direction and magnitude.

**Conclusions:** In this exploratory cross-sectional analysis, sedentary-time modification of the association between DXA-derived leg lean mass and leg BMD varied according to sex and age. The sex difference was small at younger ages and became progressively more apparent with increasing age. These findings suggest that behavioral and demographic context may contribute to variation in the muscle-bone relationship itself, although confirmation in independent and longitudinal datasets is needed.

## Introduction

Skeletal muscle and bone are functionally and biologically interconnected components of the musculoskeletal system [1,2]. Mechanical loading generated by muscle contraction contributes to bone maintenance and adaptation, while muscle and bone may also interact through endocrine and paracrine pathways [1–3]. Consistent with this concept, lean mass is positively associated with bone mineral density (BMD) across skeletal sites and populations [4,5]. A meta-analysis of 44 studies involving more than 20,000 adults found a positive association between lean mass and BMD, with some variation according to sex and menopausal status [4]. These observations suggest that musculoskeletal health may be characterized not only by the amount of muscle or bone considered separately, but also by the relationship between these tissues.

Both lean mass and BMD change across adulthood, and their age-related patterns differ between men and women [5–9]. Evidence also suggests that muscle-bone relationships may differ according to age and sex [5–8]. A recent study across a wide adult age range found positive associations of lower-body lean mass and strength with hip BMD, while also identifying age- and sex-related differences across musculoskeletal measures [8]. Such findings raise the possibility that the association between regional lean mass and bone may not represent a fixed relationship across adulthood. Behavioral factors that alter mechanical loading or musculoskeletal use could therefore plausibly modify this relationship [1,3].

Sedentary behavior is a potentially modifiable behavioral factor relevant to musculoskeletal health [10–15]. Previous population-based studies, including analyses of the National Health and Nutrition Examination Survey (NHANES), have examined associations between sedentary behavior, physical activity, and BMD [10–16]. However, these associations have not been uniform across skeletal sites, age groups, or sex [10–15]. For example, objectively measured sedentary behavior was associated with lower femoral BMD in women but not in men in an NHANES analysis [10], whereas other NHANES studies have reported age-dependent, sex-dependent, or indirect associations between sedentary activity and BMD [11,12]. More recent analyses likewise suggest that associations of sedentary time or physical activity with BMD depend on behavioral, demographic, and anatomical context [12–15]. Across these studies, sedentary behavior has generally been examined as an exposure associated with BMD, rather than as a modifier of the relationship between lean mass and BMD [10–16].

Accordingly, publicly available NHANES 2017–2018 data were used to explore whether sedentary time modifies the association between DXA-derived leg lean mass and leg BMD in U.S. adults aged 18–59 years, and whether this modification varies according to sex and age. Whether sedentary time modifies the lean mass–BMD association itself, and whether such modification differs by sex and age, remains poorly characterized [8,10–16]. The nationally representative NHANES dataset, which combines regional DXA measurements with behavioral and demographic information, provides an opportunity to examine such higher-order effect-modification patterns within an existing population-based resource [17–19]. Given the exploratory nature of the analysis, the objective was to characterize the pattern and magnitude of these associations rather than to test a prespecified causal hypothesis.

## Methods

### Study population and data source

This exploratory cross-sectional study used data from the 2017–2018 cycle of the U.S. National Health and Nutrition Examination Survey (NHANES), a complex multistage probability survey designed to generate nationally representative estimates for the U.S. civilian noninstitutionalized population [17].

The present analysis was restricted to adults aged 18–59 years. The frozen analysis dataset retained 3,706 participants in this age range before model-specific complete-case exclusions. The primary regression analysis included 2,543 participants with complete data for bilateral leg lean mass, bilateral leg BMD, sedentary time, age, sex, race/ethnicity, height, and the required NHANES survey-design variables.

### DXA-derived leg lean mass and bone mineral density

Whole-body dual-energy X-ray absorptiometry (DXA) measurements were used to derive the primary exposure and outcome variables [18]. Left and right leg lean mass excluding bone mineral content were obtained from the NHANES DXA variables DXDLLLE and DXDRLLE, respectively [18]. Mean bilateral leg lean mass was calculated as the mean of the left- and right-leg values and converted from grams to kilograms. Left and right leg BMD were obtained from DXXLLBMD and DXXRLBMD, respectively [18], and mean bilateral leg BMD was calculated as their arithmetic mean (g/cm^2^). DXA-derived leg lean mass was treated as a proxy for lower-limb muscle mass rather than a direct measure of skeletal muscle mass [20].

### Sedentary time

Daily sedentary time was obtained from PAD680 in the NHANES Physical Activity Questionnaire, which is based on the Global Physical Activity Questionnaire (GPAQ) and was administered by trained interviewers using computer-assisted personal interviewing [19]. PAD680 assesses the time usually spent sitting on a typical day, excluding sleep [19]. Sedentary time was converted from minutes per day to hours per day; NHANES special response values 7777 and 9999 were treated as missing [19]. Sedentary time was analyzed as a continuous variable. Lower sedentary time was not interpreted as equivalent to higher physical activity.

### Covariates and effect modifiers

Age was analyzed as a continuous variable in the primary interaction model. Sex was defined using RIAGENDR (male or female). Race/ethnicity was represented by RIDRETH3 and height by BMXHT. Sex and age were examined as effect modifiers rather than solely as adjustment covariates. The primary model was intentionally parsimonious and adjusted for race/ethnicity and height.

### Statistical analysis

All primary inferential analyses accounted for the complex NHANES survey design using the 2-year Mobile Examination Center examination weight (WTMEC2YR), masked variance stratum (SDMVSTRA), and primary sampling unit (SDMVPSU) [17]. The analytic sample contained 15 strata and 30 nested primary sampling units, corresponding to 15 design degrees of freedom.

Descriptive analyses examined age-related patterns in mean bilateral leg lean mass, mean bilateral leg BMD, and sedentary time according to sex. The observed relationship between leg lean mass and BMD was additionally examined using sex-specific scatterplots and locally weighted smooth curves.

Descriptive weighted linear and quadratic models were used to assess the degree of curvature in this relationship.

The main inferential analysis used a survey-weighted regression model of mean bilateral leg BMD including leg lean mass, sedentary time, sex, continuous age, and their four-way interaction, with adjustment for race/ethnicity and height. The coefficient for the leg lean mass × sedentary time × sex × age term quantified whether the sex difference in sedentary-time modification of the lean mass–BMD association changed continuously with age. Conditional leg lean mass × sedentary time interaction estimates were calculated at representative ages of 25, 35, 45, and 55 years for interpretation.

Because visual inspection suggested modest nonlinearity in the lean mass-BMD association, sensitivity analyses added a quadratic leg lean-mass term and, in a second specification, allowed the quadratic term to differ by sex. These analyses were used to assess the stability of the direction and magnitude of the four-way interaction rather than to search for an alternative statistically significant model. All analyses were exploratory; no primary hypothesis or significance threshold was prespecified before examination of the data.

### Use of artificial intelligence-assisted tools

OpenAI ChatGPT was used as an AI-assisted tool during development of analysis code, preparation of figures and tables, and manuscript drafting and editing. All analytical decisions, statistical results, source-data verification, interpretation of findings, and final manuscript content were reviewed and approved by the author, who takes full responsibility for the accuracy and integrity of the work.

## Results

### Participant characteristics and age-related patterns

The primary complete-case survey-weighted analysis included 2,543 adults aged 18-59 years, comprising 1,191 men and 1,352 women.

**Table 1.**
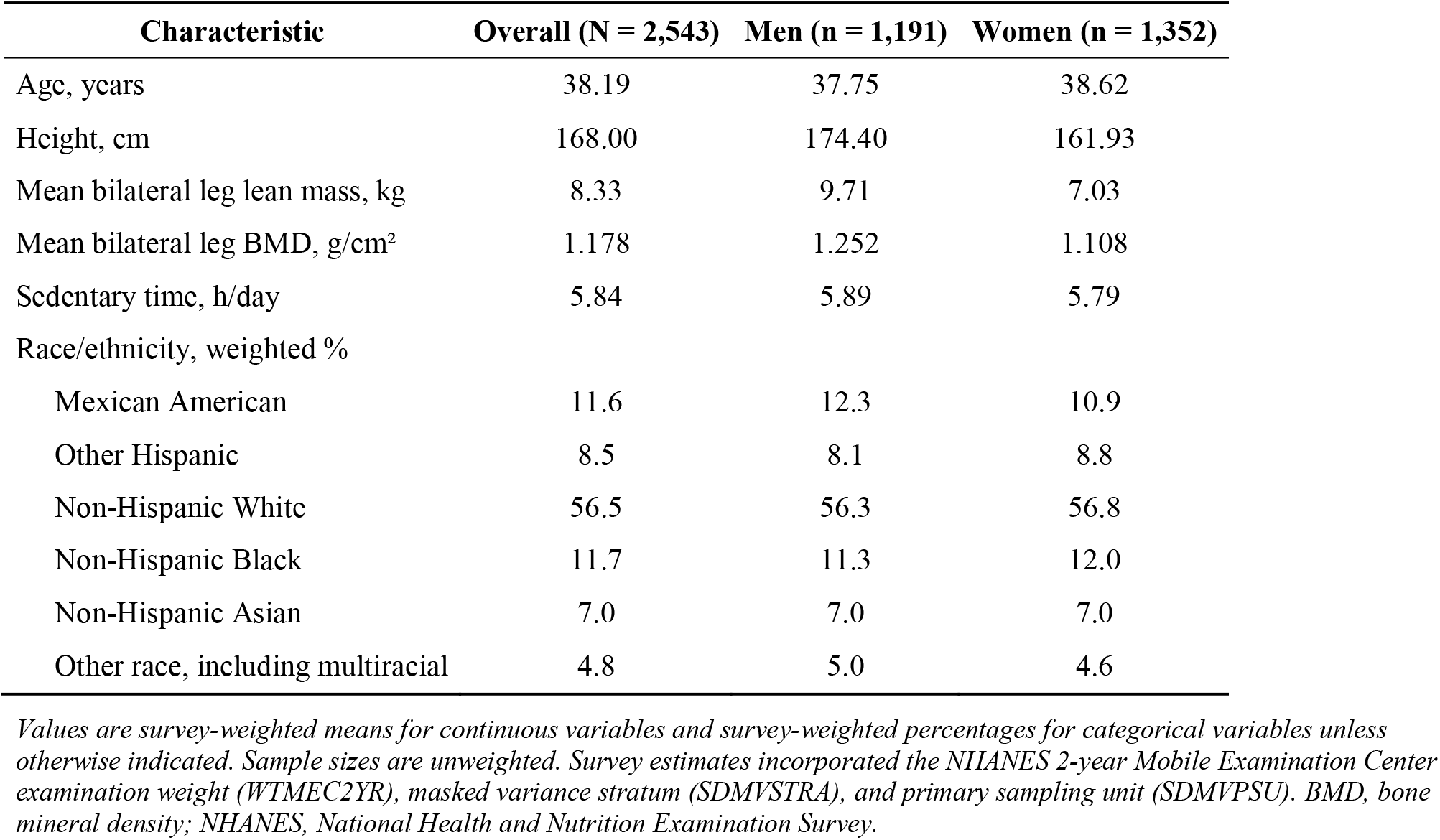
Characteristics of the analytic sample according to sex.

| Characteristic | Overall (N = 2,543) | Men (n = 1,191) | Women (n = 1,352) |
| --- | --- | --- | --- |
| Age, years | 38.19 | 37.75 | 38.62 |
| Height, cm | 168.00 | 174.40 | 161.93 |
| Mean bilateral leg lean mass, kg | 8.33 | 9.71 | 7.03 |
| Mean bilateral leg BMD, g/cm<sup>2</sup> | 1.178 | 1.252 | 1.108 |
| Sedentary time, h/day | 5.84 | 5.89 | 5.79 |
| Race/ethnicity, weighted % |  |  |  |
| Mexican American | 11.6 | 12.3 | 10.9 |
| Other Hispanic | 8.5 | 8.1 | 8.8 |
| Non-Hispanic White | 56.5 | 56.3 | 56.8 |
| Non-Hispanic Black | 11.7 | 11.3 | 12.0 |
| Non-Hispanic Asian | 7.0 | 7.0 | 7.0 |
| Other race, including multiracial | 4.8 | 5.0 | 4.6 |

*Values are survey-weighted means for continuous variables and survey-weighted percentages for categorical variables unless otherwise indicated. Sample sizes are unweighted. Survey estimates incorporated the NHANES 2-year Mobile Examination Center examination weight (WTMEC2YR), masked variance stratum (SDMVSTRA), and primary sampling unit (SDMVPSU). BMD, bone mineral density; NHANES, National Health and Nutrition Examination Survey.*

Age-related patterns in mean bilateral leg lean mass, mean bilateral leg BMD, and sedentary time are shown in **Figure 1**.

**Figure 1.**
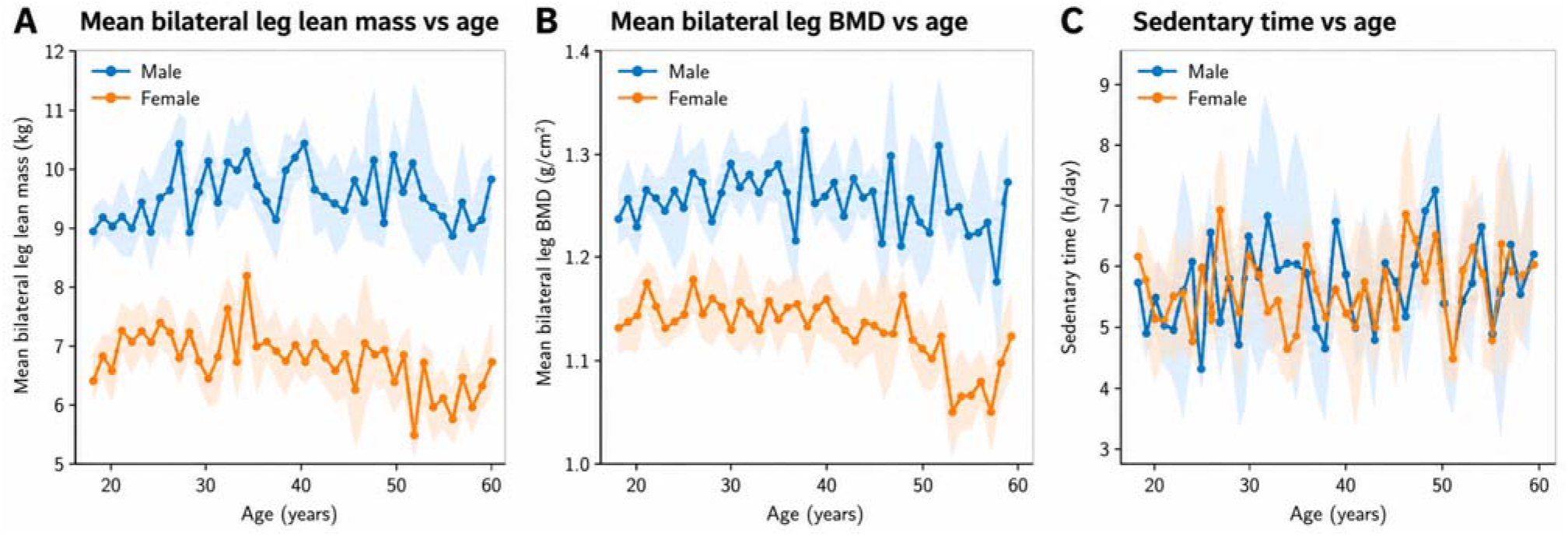
Age-related patterns in leg lean mass, leg bone mineral density, and sedentary time according to sex. Survey-weighted mean values of (A) mean bilateral leg lean mass, (B) mean bilateral leg bone mineral density (BMD), and (C) daily sedentary time are shown across ages 18–59 years for men and women. Shaded areas indicate 95% confidence intervals.

Leg lean mass was consistently greater in men than in women across the age range. Survey-weighted mean leg lean mass in men was approximately 9.45, 10.06, 9.98, and 9.62 kg at ages 18-29, 30-39, 40-49, and 50-59 years, respectively. Corresponding values in women were 7.18, 7.34, 7.12, and 6.53 kg, indicating a more apparent decline among women in the oldest age group.

Mean leg BMD in men was approximately 1.249, 1.271, 1.255, and 1.238 g/cm^2^ across the four age groups. In women, the corresponding values were 1.127, 1.127, 1.120, and 1.061 g/cm^2^, with a more pronounced decline at ages 50-59 years.

In contrast, sedentary time did not show a corresponding monotonic increase with age. Mean sedentary time in men was approximately 5.45, 6.15, 5.98, and 5.91 h/day across the four age groups, compared with 5.64, 5.63, 6.01, and 5.79 h/day in women. Thus, the age-related changes in leg lean mass and BMD, particularly among women, were not accompanied by a parallel increase in mean sedentary time.

### Association between leg lean mass and BMD

Mean bilateral leg lean mass was positively associated with mean bilateral leg BMD in both men and women (**Figure 2**). Visual inspection of the observed data and locally weighted smooth curves indicated that the association was broadly positive across the main range of leg lean mass, although modest nonlinearity was evident, particularly among women.

**Figure 2.**
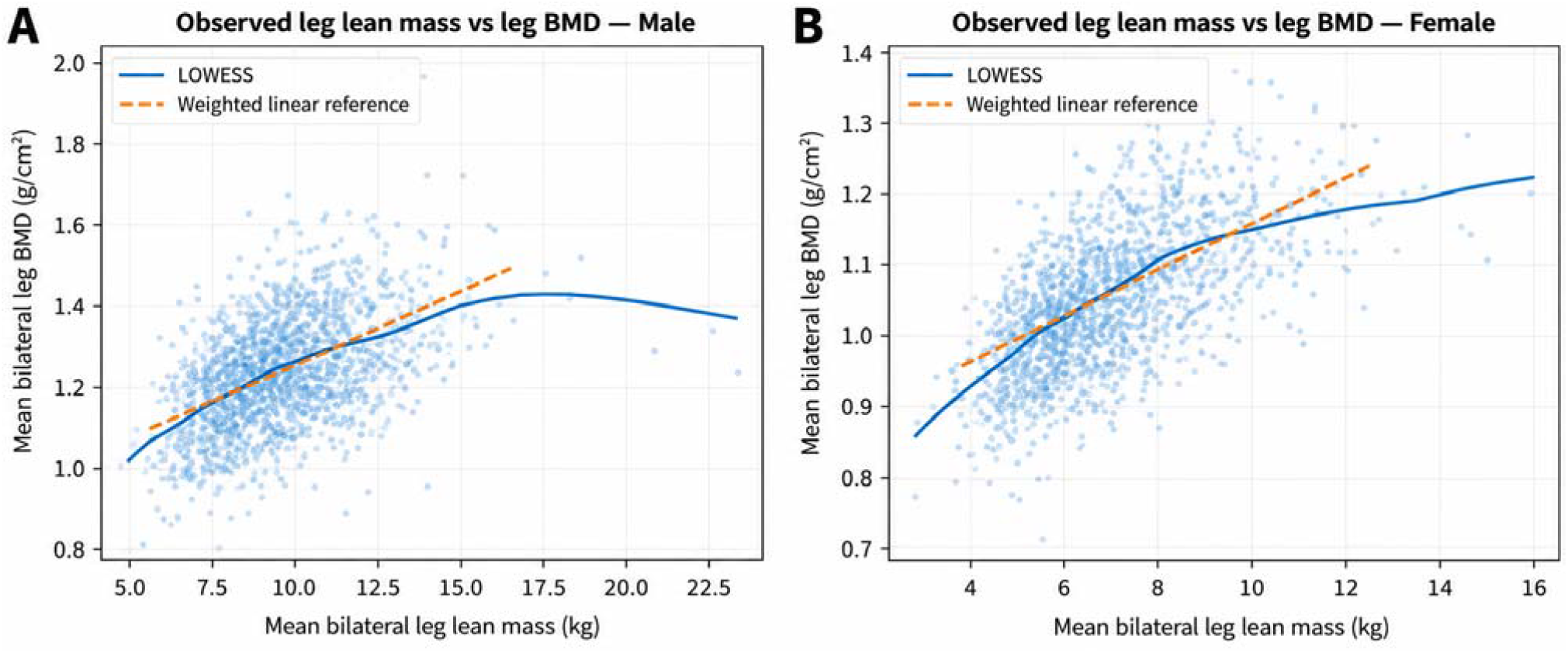
Observed association between leg lean mass and bone mineral density according to sex. Scatterplots show the observed relationship between mean bilateral leg lean mass and mean bilateral leg bone mineral density (BMD) in (A) men and (B) women. Solid curves represent locally weighted scatterplot smoothing (LOWESS), and dashed lines represent weighted linear regression fits.

In descriptive weighted models, a simple linear specification accounted for approximately 19.8% of the variation in leg BMD among men and 35.4% among women. Addition of a quadratic lean-mass term increased R^2^ from 0.198 to 0.207 in men and from 0.354 to 0.384 in women, supporting the presence of some curvature in the lean mass-BMD relationship.

### Sex differences in sedentary-time modification of the lean mass-BMD association

In the survey-weighted model including leg lean mass × sedentary time × sex, the lean mass-by-sedentary time interaction was negative but imprecisely estimated in men (β = −0.000853 g/cm^2^ per kg per h/day, 95% CI −0.002675 to 0.000968; P = 0.334) and positive in women (β = 0.000813, 95% CI 0.000158 to 0.001468; P = 0.018). The three-way leg lean mass × sedentary time × sex interaction was β = 0.001666 (95% CI 0.000232 to 0.003100; P = 0.026), indicating a sex difference in the modification of the lean mass-BMD association by sedentary time.

### Age-dependent sex differences in effect modification

Whether this sex difference varied continuously with age was then examined (**Figure 3**). In the survey-weighted four-way model, the leg lean mass × sedentary time × sex × age interaction was positive (β = 0.000111 per year, 95% CI 0.000039 to 0.000183; P = 0.0051). This estimate indicates that the sex difference in sedentary-time modification of the lean mass-BMD association increased with age.

**Figure 3.**
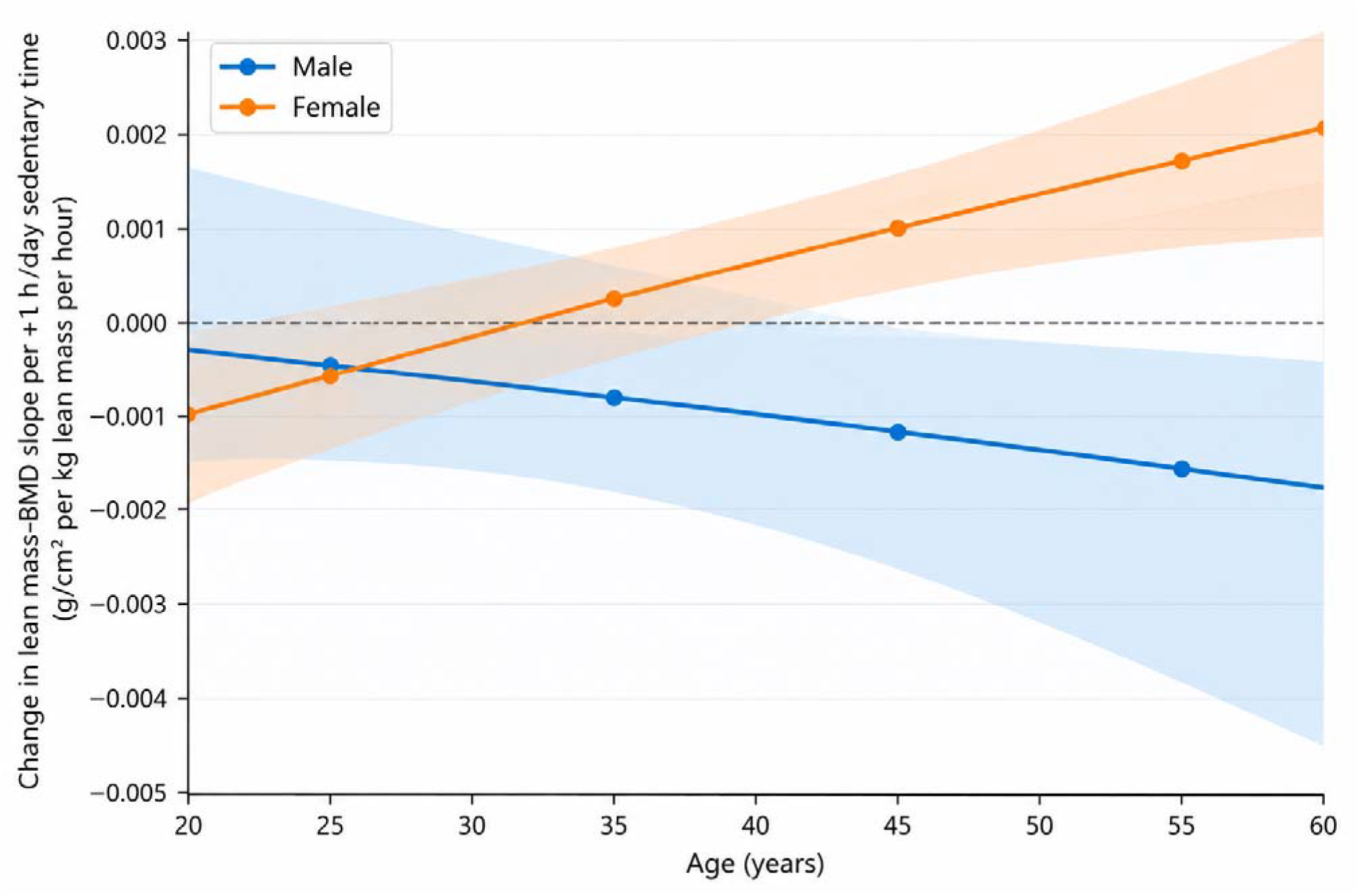
Age-dependent sex differences in sedentary-time modification of the association between leg lean mass and bone mineral density. Curves represent the estimated change in the leg lean mass– BMD slope associated with a 1-h/day increase in sedentary time, with shaded areas indicating 95% confidence intervals. Positive values indicate strengthening and negative values indicate weakening of the lean mass–BMD association with increasing sedentary time. Points indicate conditional estimates at ages 25, 35, 45, and 55 years.

Conditional estimates at representative ages of 25, 35, 45, and 55 years are shown in **Table S1**. At age 25 years, the estimated leg lean mass × sedentary time interaction was similar in men (β = −0.000475; P = 0.597) and women (β = −0.000550; P = 0.279), with little evidence of a sex difference. At age 35 years, estimates began to diverge (men, β = −0.000819; P = 0.348; women, β = 0.000217; P = 0.575), although the sex difference remained uncertain (P = 0.183).

At age 45 years, the estimated interaction remained negative but imprecise in men (β = −0.001164; P = 0.264) and was positive in women (β = 0.000985; P = 0.014), with a conditional sex-difference P value of 0.027. At age 55 years, the corresponding estimates were −0.001509 in men (P = 0.258) and 0.001752 in women (P = 0.0010), with a conditional sex-difference P value of 0.0097. Overall, estimates were similar between men and women at younger ages but progressively diverged with increasing age. Female estimates became increasingly positive, whereas male estimates tended toward negative values but remained imprecisely estimated.

### Sensitivity analysis for nonlinearity

Sensitivity analyses incorporating quadratic specifications of leg lean mass are summarized in **Table S2**. Because **Figure 2** suggested modest nonlinearity, particularly among women, the robustness of the four-way interaction to quadratic specifications of leg lean mass was examined. Addition of a quadratic lean-mass term attenuated the four-way interaction estimate by approximately 9% (β = 0.000101; P = 0.055). Allowing the quadratic term to differ by sex yielded a similar estimate (β = 0.000098; P = 0.048), representing approximately 12% attenuation relative to the primary linear model. Thus, the direction and magnitude of the age-dependent sex difference were broadly preserved, although statistical precision was reduced under nonlinear specifications.

## Discussion

In this exploratory analysis of nationally representative U.S. adults, the association between DXA-derived leg lean mass and leg BMD varied according to sedentary time, and the pattern of this modification differed by sex and age. The sex difference was small at younger ages but became progressively more apparent with increasing age. In women, the estimated modification by sedentary time became increasingly positive at older ages, whereas estimates in men tended in the opposite direction but remained imprecise. Importantly, these patterns should not be interpreted as evidence that greater sedentary time is beneficial for bone in women or detrimental in men. Rather, they indicate that the cross-sectional association between leg lean mass and BMD may not be constant across behavioral and demographic contexts.

The positive association between lean mass and BMD observed in the present study is consistent with extensive previous evidence supporting functional and biological interactions between muscle and bone [1–6,9]. Both mechanical loading and shared endocrine or paracrine pathways have been proposed to contribute to this relationship [1–3]. Recent evidence further indicates that lower-body lean mass and strength are positively associated with BMD across adulthood, with age- and sex-related variation in musculoskeletal measures [7,8]. The present findings extend this perspective by suggesting that sedentary time may represent an additional dimension along which the lean mass–BMD association varies. Previous studies using NHANES and other population-based datasets have examined direct associations of sedentary behavior with BMD, with findings differing according to sex, age, skeletal site, and measurement method [10–16]. The present results may therefore help explain why associations between sedentary behavior and bone outcomes have not been uniform across populations: sedentary time may relate not only to BMD itself but also to the relationship between muscle-related and bone-related phenotypes.

The age-related pattern observed in women deserves particular consideration. In descriptive analyses, both leg lean mass and leg BMD were lower among women aged 50–59 years than among younger women, whereas mean sedentary time did not show a corresponding increase. Thus, the age-dependent divergence observed in the interaction model cannot be explained simply by older women reporting more sedentary time. Midlife in women is accompanied by substantial changes in bone mass, body composition, and sex-hormone environment, particularly across the menopausal transition [21–25]. Rapid bone loss occurs around the menopausal transition, while lean mass also changes across this period [21–24]. Changes in muscle quantity and function associated with aging and the menopausal transition may plausibly alter the relationship between measured lean mass and its functional effect on bone [23–25]. These observations provide biologically plausible hypotheses for an age-dependent change in the muscle–bone relationship. However, menopausal status, circulating sex hormones, muscle strength, and muscle quality were not incorporated into the present analysis. The observed age pattern should therefore not be interpreted as evidence of a menopausal mechanism.

The opposite directions of the estimated sedentary-time modification in older men and women are more difficult to interpret. In particular, a positive interaction in women does not imply that sedentary behavior improves skeletal health. An interaction coefficient describes how the cross-sectional slope relating lean mass to BMD differs across levels of sedentary time; it does not describe the effect of sedentary time on BMD itself. Several processes could produce such a pattern, including sex-specific changes in muscle quantity or function, mechanical loading, body composition, or the relative rates at which muscle and bone change with age [1,3,23–25]. Differential residual confounding or selection may also contribute. Because lean mass measured by DXA represents lean soft tissue rather than skeletal muscle mass directly, the biological meaning of a given kilogram of leg lean mass may also differ according to sex and age [20]. These possibilities cannot be distinguished using the present cross-sectional analysis and should be considered hypotheses for future investigation rather than explanations established by the findings.

A further strength of this study is the use of a publicly available, nationally representative dataset containing regional DXA, behavioral, and demographic measurements within the same participants [17]. Large public resources such as NHANES permit existing measurements to be recombined to address questions beyond the original focus of data collection. In the present study, this allowed examination of not only established associations among lean mass, BMD, and sedentary behavior separately, but also whether the structure of the lean mass–BMD association varied across behavioral and demographic contexts. Such secondary analyses are particularly useful for identifying interaction patterns that can subsequently be tested in longitudinal or purpose-designed studies. The use of public data also facilitates independent reproduction and alternative model specifications, which is particularly important for exploratory higher-order interaction analyses.

The findings should nevertheless be interpreted cautiously. First, NHANES is cross-sectional, and temporal or causal relationships among sedentary time, lean mass, and BMD cannot be established. Second, sedentary time was self-reported and may be affected by recall or reporting error [26,27].

Third, DXA-derived leg lean mass is a measure of lean soft tissue and only a proxy for skeletal muscle mass [20]; muscle strength and muscle quality were not assessed in the primary analysis. Fourth, the four-way interaction emerged from an exploratory analytical sequence rather than from a prespecified confirmatory hypothesis. Although the analysis incorporated NHANES sampling weights, strata, and primary sampling units, and the direction and approximate magnitude of the four-way interaction were retained after incorporating quadratic lean-mass terms, statistical precision was reduced in these sensitivity models. The primary estimate should therefore not be regarded as definitive evidence solely on the basis of its P value. Residual confounding by factors not included in the present parsimonious model also remains possible.

Finally, the present findings suggest a potentially useful direction for future research. Muscle mass and BMD are commonly evaluated as separate musculoskeletal characteristics, but the relationship between them may itself contain information about musculoskeletal status. If the age- and sex-dependent modification observed here is reproduced in independent datasets, future studies could examine the lean mass–BMD relationship as a phenotype in its own right and determine whether its variation predicts subsequent bone loss, fracture, functional decline, or other musculoskeletal outcomes. Longitudinal studies incorporating objectively measured sedentary behavior, muscle strength and quality, menopausal status, and hormonal measurements would be particularly informative. The present analysis should therefore be viewed primarily as hypothesis-generating evidence that the muscle–bone relationship may vary systematically according to behavioral and demographic context.

## Conclusions

In this exploratory cross-sectional analysis of U.S. adults, sedentary-time modification of the association between DXA-derived leg lean mass and leg BMD differed according to sex and changed with age. The sex difference was limited at younger ages and became more apparent in later adulthood, without a parallel age-related increase in sedentary time itself. These findings suggest that the muscle–bone relationship may vary across behavioral and demographic contexts, but independent and longitudinal studies are needed to determine the biological and clinical significance of this pattern.

## Data Availability

The study used only publicly available, de-identified data from the National Health and Nutrition Examination Survey (NHANES) 2017-2018 that were openly available before initiation of the present study. Data were obtained from the publicly accessible NHANES 2017-2018 data files provided by the National Center for Health Statistics, including the Demographic Variables and Sample Weights (DEMO_J), Dual-Energy X-ray Absorptiometry - Whole Body (DXX_J), and Physical Activity (PAQ_J) datasets. Links to the original data and documentation pages are provided below.

https://wwwn.cdc.gov/nchs/nhanes/continuousnhanes/default.aspx?BeginYear=2017

https://wwwn.cdc.gov/Nchs/Data/Nhanes/Public/2017/DataFiles/DEMO_J.htm

https://wwwn.cdc.gov/Nchs/Data/Nhanes/Public/2017/DataFiles/DXX_J.htm

https://wwwn.cdc.gov/Nchs/Data/Nhanes/Public/2017/DataFiles/PAQ_J.htm

## Declarations

### Ethics approval and consent to participate

This study was a secondary analysis of publicly available, de-identified data from the National Health and Nutrition Examination Survey (NHANES). The original NHANES protocols were approved by the National Center for Health Statistics Research Ethics Review Board, and written informed consent was obtained from all participants. No additional institutional review board approval or informed consent was required for the present secondary analysis of publicly available de-identified data.

### Competing interests

The author declares no competing interests.

### Funding

This research was funded by Japan Society for the Promotion of Science (JSPS) KAKENHI, grant number 22K06618 to Y.K.

### Data and code availability

The original NHANES 2017–2018 data used in this study are publicly available from the National Center for Health Statistics. The derived analysis dataset, data dictionary, analysis code, figure and table source data, and supporting documentation will be made publicly available through Zenodo at the time of preprint posting (doi:10.5281/zenodo.22734947).

### Author contributions

Y.K. conceived the study, developed the research question and analytical strategy, interpreted the data, and drafted and critically revised the manuscript. Y.K. takes full responsibility for the integrity of the work and approved the final manuscript.

## Notes

### Competing Interest Statement

The authors have declared no competing interest.

### Author Declarations

The study used only publicly available, de-identified data from the National Health and Nutrition Examination Survey (NHANES) 2017-2018, which were openly available before initiation of the present study. The original datasets, documentation, and codebooks are publicly available from the National Center for Health Statistics (NCHS), Centers for Disease Control and Prevention (CDC), through the NHANES 2017-2018 data portal.

