## Supplemental Tables S1 and S2 for "Age and sex differences in the modification of the leg lean mass–bone mineral density association by sedentary time in U.S. adults"

**Table S1. Conditional sedentary-time modification of the leg lean mass–BMD association at representative ages according to sex**

| Age, years | Men, β (95% CI) | P value | Women, β (95% CI) | P value | P for sex difference |
|---|---|---|---|---|---|
| 25 | −0.000475 (−0.002349 to 0.001399) | 0.597 | −0.000550 (−0.001594 to 0.000494) | 0.279 | 0.922 |
| 35 | −0.000819 (−0.002621 to 0.000983) | 0.348 | 0.000217 (−0.000590 to 0.001024) | 0.575 | 0.183 |
| 45 | −0.001164 (−0.003302 to 0.000974) | 0.264 | 0.000985 (0.000230 to 0.001740) | 0.014 | 0.027 |
| 55 | −0.001509 (−0.004244 to 0.001226) | 0.258 | 0.001752 (0.000835 to 0.002669) | 0.0010 | 0.0097 |

*β represents the estimated change in the leg lean mass–BMD slope associated with a 1-h/day increase in sedentary time (g/cm² per kg lean mass per hour). Estimates were derived from the survey-weighted model including the leg lean mass × sedentary time × sex × continuous age interaction, with adjustment for race/ethnicity and height. Positive values indicate strengthening and negative values indicate weakening of the leg lean mass–BMD association with increasing sedentary time. P values for sex difference compare the conditional interaction estimates between men and women at each age.*

**Table S2. Sensitivity analyses for nonlinearity in the leg lean mass–BMD association**

| Model | Four-way interaction β (95% CI) | P value | Change from primary estimate |
|---|---|---|---|
| Primary linear model | 0.000111 (0.000039 to 0.000183) | 0.0051 | Reference |
| + quadratic leg lean mass term | 0.000101 (−0.000001 to 0.000203) | 0.055 | −9% |
| + quadratic leg lean mass term and lean mass² × sex | 0.000098 (0.000000 to 0.000196) | 0.048 | −12% |
